# The neurophysiological brain-fingerprint of Parkinson’s disease

**DOI:** 10.1101/2023.02.03.23285441

**Authors:** Jason da Silva Castanheira, Alex I. Wiesman, Justine Y. Hansen, Bratislav Misic, Sylvain Baillet, PREVENT-AD Research Group, Network Quebec Parkinson

**Affiliations:** Montreal Neurological Institute, McGill University, Montreal QC, Canada

**Keywords:** Movement disorders, Parkinson’s disease, neural dynamics, oscillations, arrhythmic brain activity, magnetoencephalography, brain-fingerprinting

## Abstract

In this study, we investigate the clinical potential of brain-fingerprints derived from electrophysiological brain activity for diagnostics and progression monitoring of Parkinson’s disease (PD). We obtained brain-fingerprints from PD patients and age-matched healthy controls using short, task-free magnetoencephalographic recordings. The rhythmic components of the individual brain-fingerprint distinguished between patients and healthy participants with approximately 90% accuracy. The most prominent cortical features of the Parkinson’s brain-fingerprint mapped to polyrhythmic activity in unimodal sensorimotor regions. Leveraging these features, we also show that Parkinson’s disease stages can be decoded directly from cortical neurophysiological activity. Additionally, our study reveals that the cortical topography of the Parkinson’s brain-fingerprint aligns with that of neurotransmitter systems affected by the disease’s pathophysiology. We further demonstrate that the arrhythmic components of cortical activity are more variable over short periods of time in patients with Parkinson’s disease than in healthy controls, making individual differentiation between patients based on these features more challenging and explaining previous negative published results. Overall, we outline patient-specific rhythmic brain signaling features that provide insights into both the neurophysiological signature and clinical staging of Parkinson’s disease. For this reason, the proposed definition of a rhythmic brain-fingerprint of Parkinson’s disease may contribute to novel, refined approaches to patient stratification and to the improved identification and testing of therapeutic neurostimulation targets.

**Lay summary:** We propose a new method to help diagnose and monitor Parkinson’s disease (PD) using patients’ unique *brain-fingerprint*. These fingerprints are based on the brain’s electrical activity, which we measured without any specific tasks, using a technique called magnetoencephalography. Remarkably, we found that these brain-fingerprints can differentiate between people with Parkinson’s and those without, with about 90% accuracy. Specifically, we noticed that certain rhythmic patterns in the brain, particularly in areas involved in sensory and motor functions, were key indicators of Parkinson’s. Interestingly, these patterns also helped us identify the different stages of the disease.

Additionally, our research shows that the arrangement of these brain-fingerprints in Parkinson’s patients corresponds to how the neurochemistry of the brain is impacted by the disease. We also observed that certain irregular patterns in the brain’s activity, which vary more from moment to moment in Parkinson’s patients, make it harder to distinguish between individuals based on these features alone. This finding sheds light on why previous studies reported challenges with similar approaches.

Overall, our study offers new insights into the unique brain activity patterns in Parkinson’s disease and suggests that individual brain-fingerprints could be valuable in tailoring treatment plans and developing new therapies for this condition.

## Introduction

The neurophysiological underpinnings of Parkinson’s disease (PD) are characterized by a spectrum of motor and non-motor symptoms that vary widely among patients, and their fundamental nature continues to be a subject of extensive research^1–3^. This variation in symptoms is paralleled by PD’s diverse structural alterations^4–7^, along with changes in hemodynamic and electrophysiological brain activity compared to healthy individuals^1,8–12^. Notably, electrophysiological changes in PD concern both the rhythmic and arrhythmic components of neurophysiological signals^8,9,12–15^. Brain-network characteristics, as highlighted in previous studies using functional connectome analysis with functional magnetic resonance imaging (fMRI) and other brain mapping techniques, also deviate from those in health and correlate with PD’s hallmark motor and cognitive impairments^16–19^.

Recent methodological advances have employed fMRI connectomes to derive *brain-fingerprints*, providing biometric differentiation based on individual neuroimaging phenotypes^20–23^. This concept posits that the neuroimaging phenotypes of an individual remain relatively stable over time, forming the basis for distinctive brain-fingerprints^20,21,24^. Such brain-fingerprinting has enabled exploration of the neurophysiological bases of complex traits and behaviors in healthy subjects_20,21,23–27._

However, subsequent studies have indicated an increased variability in brain-fingerprints over time in PD^26,28,29^, which challenges the differentiation between patients on the basis of this neuroimaging phenotype. For example, a recent study with magnetoencephalography (MEG) showed that the differentiation accuracy between the brain-fingerprints of patients with PD, derived from connectomes, declines with the severity of their motor symptoms^30^. This also challenges the ability to distinguish PD patients from healthy controls based on brain-fingerprints, thereby raising questions about the relevance and effectiveness of these approaches in neurological disease research, or for testing novel therapeutic pathways.

A plausible explanation for the apparent limited impact of brain-fingerprinting to clinical research so far may be an inherent instability in the brain activity of PD patients over short periods. For instance, previous work has shown that hemodynamic signals from functional near-infrared spectroscopy (fNIRS) are more variable in patients with severe PD symptoms^31^. This suggests that electrophysiological activity in PD is also likely to exhibit greater temporal variability, especially in brain regions with strong coupling between electrophysiological and hemodynamic signals^32^.

This short-term variability within patients challenges the definition of a stable brain-fingerprint profile that would accurately characterize an individual’s disease stage. Further research is needed to determine whether this increased variability affects the entire frequency spectrum of electrophysiological activity or is confined to specific rhythmic or arrhythmic components^33,34^.

In a recent study with healthy young adult participants, we showed that frequency-specific measures of electrophysiological activity across the cortex, derived from brief, task-free MEG data, define *spectral* brain-fingerprints that are specific to each individual over remarkably prolonged periods of time^24^.

In the present study, we extend this approach and confirm that the electrophysiological brain-fingerprint of patients with PD exhibits greater variability over time compared to that of healthy controls. However, this variability is predominantly driven by the arrhythmic component of the neurophysiological power spectrum. In contrast, rhythmic features of the PD brain-fingerprint remain remarkably stable, enabling effective differentiation between PD patients and healthy controls, and among patients themselves. We highlight the clinical significance of these stable features by relating them to individual disease stages, and their cortical topography to the functional hierarchy of the cortex^35^ and atlas maps of cortical neurotransmitter systems relevant to PD neuropathophysiology^36^.

## Methods

### Participants

Participants for this study were selected from a diverse age group (40-82 years) and included healthy controls as well as patients with mild to moderate idiopathic Parkinson’s Disease (PD). We aggregated data from multiple sources. We utilized data from 79 PD patients who were part of the Quebec Parkinson Network (QPN; https://rpq-qpn.ca^84^). These patients had undergone extensive clinical, neurophysiological, and biological profiling. All enrolled patients in the QPN study were on a stable dose of antiparkinsonian medication and demonstrated satisfactory clinical responses. They were instructed to continue their medication regimen as prescribed before any data collection. We included data from QPN participants who had complete and usable Magnetoencephalography (MEG; 275 channels whole-head CTF; Port Coquitlam, British Columbia, Canada) clinical, and demographic data.

Our main control group comprised demographically matched participants from the PREVENT-AD (N=50)^85^ and OMEGA (N=4)^86^ studies, ensuring a comparison group that mirrors the age and demographic characteristics of the PD group. We replicated our observations using a second sample of healthy controls from the Cambridge Center for Aging Neuroscience (CamCAN) dataset (N= 370 healthy adults, 40-78 years old, 58.67, SD= 11.04; 185 Females) recorded on a different MEG instrument. See “CamCAN sample of healthy controls” below for more details.

All participants underwent resting-state eyes-open MEG recordings. These recordings were conducted using a 275-channel whole-head CTF system (Port Coquitlam, British Columbia, Canada) at a sampling rate of 2400 Hz, with a 600-Hz antialiasing filter. We also applied systems built-in third-order gradient filters to the recordings. Consistency in data collection was maintained by conducting all recordings at the same site, each lasting a minimum of 10 minutes.

### Preprocessing of MEG Data

We preprocessed the MEG data using Brainstorm^87^, March-2021 distribution, on MATLAB 2019b (Mathworks, Inc., Massachusetts, USA). We adhered to established good practice guidelines^88^ and replicated the following pre-processing steps following previous published studies applied to similar data^15,89^.

We filtered the MEG sensor signals between 1–200 Hz to minimize slow-wave drifts and high-frequency noise. Then, we removed line noise artifacts at 60 Hz and harmonic frequencies, with a notch filter bank. We corrected for cardiac and ocular artifacts using Signal-Space Projectors (SSPs), derived from electro-cardiogram and electro-oculogram recordings, using an automated procedure in *Brainstorm*^87^. We segmented the MEG recordings into non-overlapping 6-second epochs and downsampled them to 600 Hz. Lastly, we screened and excluded data segments with peak-to-peak signal amplitude or maximum signal gradient exceeding ±3 absolute deviations from the median across all epochs.

### MEG source mapping

We derived brain source models from each participant’s individual T1-weighted MRI data. We segmented and labeled the MRI volumes using Freesurfer^90^. We coregistered the MEG data to these segmented MRIs using approximately 100 head points that were digitized on the day of the MEG sessions. For 14 PD patients and 3 controls who lacked usable MRI data, we warped the default Freesurfer anatomy using Brainstorm procedures to match their available head digitization points and anatomical landmarks.

We created biophysical head models for each participant using the Brainstorm overlapping-spheres model with default parameters. The MEG cortical maps consisted of 15,000 elementary dipole sources, constrained to the cortical surface, with free orientation. We computed source maps for each participant and each 6-second epoch using dynamic statistical parametric mapping (dSPM) with Brainstorm’s default parameters. To model environmental noise statistically, we processed with the same approach the two-minute empty-room recordings collected around the time of each participant’s visit.

For all epochs, we extracted individual source time series at each cortical location from the first principal component of the three elementary time courses of each triplet of elementary sources at each cortical vertex. Finally, we clustered the resulting 15,000 time series according to the Desikan-Killiany cortical parcellation^38^ into 68 regions of interest (ROIs), obtaining one representative time series per parcel from the first principal component of all source signals within each ROI.

### Derivation of spectral brain-fingerprints

We derived brain-fingerprints from the power spectrum of the ROI source time series. We calculated the Power Spectrum Density (PSD) for each parcel using Welch’s method, with a time window of 3 seconds and 50% overlap. This approach yielded PSDs in the frequency range of 0–150 Hz, at a frequency resolution of 1/3 Hz.

Each individual’s spectral brain-fingerprint was composed of the PSDs of all 68 cortical parcels, averaged across all 6-second epochs. As detailed in Results, we derived two sets of spectral brain-fingerprints based on epochs from either the first or second half of the entire MEG session recordings. We also produced spectral brain-fingerprints from shorter datasets comprising 30-second non-overlapping segments.

The feature count in each spectral brain-fingerprint totaled 68×451. We performed subsequent analyses using in-house developed code in Python (version 3.7.6) and R (version 4.2.1).

### Individual differentiation from spectral brain-fingerprints

We replicated a previously published fingerprinting approach based on the correlational differentiability of participants between data segments (as illustrated in Figure 1a-b)^24^. For each participant, we calculated all Pearson’s correlation coefficients between their first spectral brain-fingerprint and the second brain-fingerprint of every individual in the same cohort, including the participant being analyzed. The fingerprinting process per se involved a simple lookup along the rows or columns of the symmetrical interindividual correlation matrix. The highest correlation coefficient in this matrix indicated the matching participant.

**Figure 1:**
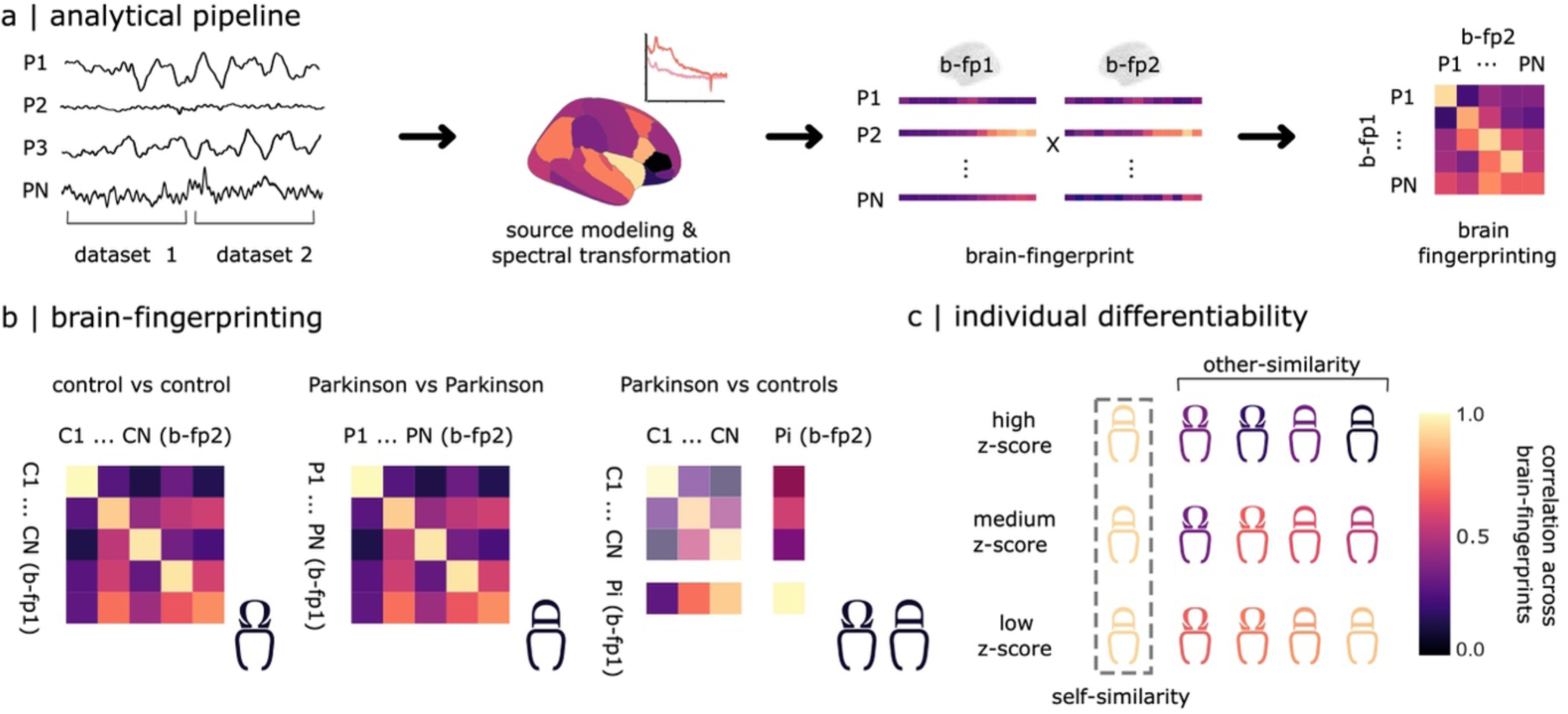
Brain-fingerprinting pipeline and study design. (a) From each participant, the power spectrum density of MEG source time series is computed for each region defined by the Desikan-Killiany atlas. This is done for two data segments (datasets 1 and 2), each containing approximately 4 minutes of clean data. The power spectra from these segments form two spectral brain-fingerprints (b-fp1 and b-fp2) for each participant ^38^. A confusion matrix, using self- and other-similarity measures of these brain-fingerprints across participants, enables inter-individual differentiation assessment. (b) We evaluated the effectiveness of this approach in differentiating individuals among three groups: i) healthy controls, ii) patients with Parkinson’s Disease (PD), and iii) each PD patient compared to healthy controls. (c) We derived an individual differentiability score for each participant, based on the self-similarity of their two brain-fingerprints. This score is z-scored against the other-similarity of their fingerprints with those of other participants in the study.

We repeated this approach for all participants in the cohort, resulting in a confusion matrix across all participants based on the two instances of their respective brain-fingerprints. We determined the overall differentiation accuracy of the brain-fingerprinting procedure by calculating the percent ratio of correctly differentiated individuals.

We addressed three types of differentiation challenges: i) differentiation among healthy participants, ii) differentiation among Parkinson’s disease patients, and iii) differentiation of each PD patient against all healthy controls (as shown in Figure 1c). Differentiating healthy participants aimed to replicate our earlier study with younger adults^24^ in an older participant group, providing a benchmark differentiation accuracy for the patient participants’ age group.

We defined individual differentiability as the ability to distinguish a participant from others in the cohort based on their brain-fingerprint. We calculated this measure as the z-scored Pearson’s correlation between the two brain-fingerprints of a given participant (self-similarity), relative to the mean and standard deviation of the correlations between this participant’s first brain-fingerprint and the second brain-fingerprints of all other participants (other-similarity).

### Bootstrapping differentiation accuracy scores

To establish confidence intervals for the average differentiation accuracy scores obtained from the fingerprinting procedure, we employed a bootstrapping method across the tested cohorts. This involved randomly selecting a subset of participants, constituting 90% of the cohort, and performing brain-fingerprinting on their data to obtain a differentiation accuracy score for that subset.

We repeated this process 1,000 times, each time with a different random subset of participants from the cohort. From the empirical distribution of these differentiation accuracies, we derived a 95% confidence interval, using the 2.5^th^ and 97.5^th^ percentiles.

### Addressing biophysical and environmental artifacts

We examined the potential impact of environmental noise and biophysical recording artifacts on the differentiation of individual participants. To do this, we correlated individual differentiability scores with the root-mean-squares (RMS) of ocular, cardiac, and head movement signals that were recorded simultaneously with MEG. These signals included data from electrocardiogram (ECG), horizontal electrooculogram (HEOG), vertical electrooculogram (VEOG), and head-coils triplet channels.

We analyzed the correlations between these three measures and the individual differentiability of each participant from the entire cohort. Additionally, we included the head motion RMS measure as a nuisance covariate in our regression model that explored the relationship between individual differentiability and motor symptoms.

To assess if environmental and instrument noise, which can vary daily, could have biased individual differentiation, we utilized the empty-room recordings collected alongside each MEG session. From these recordings, we derived pseudo brain-fingerprints for each participant, based on the cortical source maps of the noise recordings. We then calculated the differentiation accuracies from these pseudo brain-fingerprints, following the same procedure as above.

### Arrhythmic/rhythmic spectral parametrization

To evaluate the contribution of arrhythmic and rhythmic spectral components to individual differentiation, we first identified the best-fitting arrhythmic components of each individual’s brain-fingerprint spectral features in the 2-40 Hz range using *specparam* in Brainstorm. The parameters for *specparam* were set as follows: peak width limits between 0.5 and 12 Hz, a maximum of 3 peaks, a minimum peak amplitude of 3 arbitrary units (a.u.), a peak threshold of 2 standard deviations, a proximity threshold of 2 standard deviations, and a fixed aperiodic mode.

Using these arrhythmic models, we derived brain-fingerprints based solely on their features. Symmetrically, we removed the arrhythmic components from the original brain-fingerprints to isolate the rhythmic residuals and assess their contribution to inter-individual differentiation.

We then conducted the same brain-fingerprinting analyses as previously described, applying them separately to both arrhythmic and rhythmic brain-fingerprints.

### Saliency of brain-fingerprint features

We quantified the contribution of each cortical region to individual differentiation using intraclass correlations (ICC). ICC assess the agreement between two measures, in this context, indicating how consistent a particular brain-fingerprint feature is across the two brain-fingerprints of each individual compared to others in the cohort. A higher ICC for a given brain-fingerprint feature implies greater consistency across an individual’s brain-fingerprints relative to the cohort.

To illustrate the saliency of these features, we created ΔICC maps, as shown in Figure 3b and Figure S3. We first averaged the ΔICC values within each of the canonical frequency bands and then averaged these across all bands. This process involved averaging ΔICC within specified frequency ranges: delta (1–4 Hz), theta (4–8 Hz), alpha (8–13 Hz), beta (13–30 Hz), and gamma (30–50 Hz). This resulted in six ΔICC maps, one for each frequency band, which were then averaged to obtain a broadband ΔICC map.

The rationale behind this method was to give equal weight to each frequency band in the derivation of the broadband ΔICC, irrespective of their respective bandwidths. For example, while the delta band has a bandwidth of 4 Hz, the high-gamma band encompasses 100 Hz. This approach ensures a balanced representation of all frequency bands in assessing the contribution of cortical regions to individual differentiation based on brain-fingerprint features.

### CamCAN sample of healthy controls

For verifying the robustness of the ΔICC cortical map, we utilized an independent sample of healthy age-matched controls from the Cambridge Center for Aging Neuroscience (CamCAN) dataset. This data consisted of resting-state, eye-closed MEG recordings using a 306-channel VectorView MEG system (Elekta Neuromag, Helsinki). We processed the data of 370 healthy adults, aged between 40 and 78 years, from the CamCAN dataset using a pipeline similar to the one described in this paper.

We preprocessed the CamCAN dataset in a similar fashion to the other data reported here. However, we used the linearly constrained beamformer in Brainstorm with default parameters to brain map sensor data and used longer time windows of 2 seconds with a 50% overlap for power spectrum estimates of source time series.

We computed PSD estimates at each of the 68 parcels in the Desikan-Killiany atlas. We calculated ICC values for each specified frequency band. Using these ICC values, we constructed a cortical map of broadband ΔICC, following the method outlined above in Saliency of Brain-fingerprint Features.

### Computational neuroanatomy analysis

We ensured that neuroanatomical features, including those altered by Parkinson’s disease, did not influence the differentiation of participants based on their spectral brain-fingerprints. To achieve this, we measured z-scored deviations in cortical thickness for each cortical parcel in PD patients. These deviations were calculated using FreeSurfer’s recon-all, based on the mean and standard deviation of cortical thickness observed in the age-matched healthy controls.

We employed linear regression models to investigate two aspects: i) whether patients who were most differentiable based on their brain-fingerprints also exhibited greater deviations in cortical thickness, and ii) the relationship between deviations in regional cortical thickness and regional ΔICC (as depicted in Figure 3b, left panel).

### Decoding disease stage from brain-fingerprints

We used individual Hoehn & Yahr scores as markers for disease staging in PD patients^41,42^. We binarized these scores around a value of 2, creating two distinct groups to differentiate patients with unilateral symptoms from those with bilateral symptoms.

We trained a linear support vector machine (SVM) classifier in R, using default parameters. We trained a linear support vector machine (SVM) classifier in R with default parameters, to identify each patient’s disease stage category from their respective spectral brain-fingerprint features.

We conducted SVM classification for each cortical parcel independently. To train the SVM classifier, we used data from a random sample of 80% of the patients, and the remaining 20% of patients served as a test set. We recorded the percentage of these held-out patients for whom the classifier accurately identified their Hoehn & Yahr category. We repeated this classification process 1,000 times for each cortical parcel, generating an empirical distribution of disease stage classification accuracy across the cortex.

Next, we examined the spatial correlation between the cortical topographies of disease stage decoding accuracy and the regional ΔICC values from the brain-fingerprints (as discussed above in Saliency of Brain-fingerprint Features). Specifically, we correlated the differences in ICC values— subtracted between the PD-cohort fingerprinting challenge and the control-cohort challenge— across cortical ROIs with the decoding accuracies obtained from Hoehn & Yahr score decoding.

### Correspondence with cortical functional hierarchy

We examined whether the brain-fingerprints of individuals with Parkinson’s Disease (PD) align with the cortical topography of functional hierarchies in the cortex^35^. To investigate this, we focused on the spatial relationship between the ΔICC brain-fingerprint topography and the first gradient of the cortical functional hierarchy.

We calculated Pearson’s spatial correlation between the ΔICC topography of brain-fingerprints and the atlas map of the first gradient of the cortical functional hierarchy. The gradient map we used is available from *neuromaps*^36^ and was parcellated into the 68 regions of the Desikan-Killiany atlas.

To statistically evaluate the significance of these correlations, we computed Bayes factors using the *correlationBF* function in R. Additionally, we estimated p-values using permutation tests that accounted for the spatial autocorrelation inherent in the data^91,92^.

### Correlation with cortical neurotransmitter systems

Using a similar approach, we assessed the spatial correlation between the ΔICC values of brain-fingerprints and the normative atlas maps of various neurotransmitter systems. These systems were represented by maps for 19 receptors and transporters across 9 neurotransmitter systems, obtained from *neuromaps*. The neurotransmitter systems and their corresponding receptors and transporters included: Dopamine (D1, D2, DAT); Serotonin (5-HT1a, 5-HT1b, 5-HT2a, 5-HT4, 5-HT6, 5-HTT); Acetylcholine (α4β2, M1, VAChT); GABA (GABAa); Glutamate (NMDA, mGluR5); Norepinephrine (NET); Histamine (H3); Cannabinoid (CB1) and Opioid (MOR).

Each neurotransmitter system map was parcellated using the 68 regions of the Desikan-Killiany atlas. We then calculated Pearson’s spatial correlations between these neurochemical maps and the regional ΔICC values of brain-fingerprints.

To determine statistical significance, we corrected for multiple comparisons using the False Discovery Rate (FDR) method implemented in R’s *p.adjust* function^93^. We also computed Bayes factors using the *correlationBF* function in R to quantify evidence in favor of the alternative hypothesis that a spatial correlation exists.

For each significant spatial correspondence observed, we estimated p-values based on spatially constrained permutation tests^91,92^. We conducted 1,000 permutations of the neurochemical atlases using the Hungarian method. It is important to note that the reported effects might be stronger than what was observed in the spin tests from the permuted data, leading to a null p_spin_ value.

### Temporal variability of the PD brain-fingerprint

To investigate the temporal variability of PD brain-fingerprint, we utilized brain-fingerprints derived from 30-second recordings of data. This approach builds on our previous work, which demonstrated the robustness of spectral brain-fingerprints derived from brief recordings^81^.

To predict the Fisher z-transformed self-similarity (i.e., autocorrelation) of successive brain-fingerprints, we employed second-order polynomial hierarchical regression models constructed using the *lme4* package in R.

In our modeling, we nested the slope of gap duration within each subject, allowing for second-order polynomial fits for gap duration between brain fingerprints.

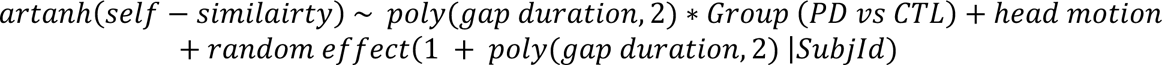

## Results

We collected at least two task-free MEG recordings, each lasting 5 minutes with participants’ eyes open, from 79 PD patients and 54 age-matched healthy controls (Prevent-AD sample; demographic details in Table S1). We then applied source-imaging to the MEG sensor data, using individual cortical surfaces derived from T1-weighted structural MRI scans^37^. For each participant, we estimated the power spectrum density (PSD) of their cortical MEG time series in the 0-150 Hz frequency range, across the cortical regions defined by the Desikan-Killiany atlas^38^. This process generated one spectral brain-fingerprint for each participant’s MEG recordings (see Methods).

Our goal with brain-fingerprinting was to quantify the distinctiveness of individual features in the brain-fingerprints of patients and healthy controls. We therefore compared the cortical spectral features from each participant’s MEG recordings with those of all other participants in our sample. By doing so, we extended our seminal brain-fingerprinting results, previously established in young healthy adults^24^, to the healthy older individuals in our present cohort. Subsequently, we applied the same analysis to differentiate PD patients. Finally, we examined if this method could reliably distinguish PD patients from their age-matched healthy counterparts (Figure 1). For the differentiation accuracy scores obtained, we calculated bootstrapped confidence intervals (CIs), as detailed in the Methods section.

### Brain-Fingerprinting Accuracy in Differentiating Healthy and PD Participants

We found that healthy participants can be differentiated from each other with 89.8% accuracy (CI [88.0, 94.0]; Figure 2a), patients with PD from each other with 77.2% accuracy (CI [74.7, 81.7]), and patients from healthy controls with 81.1% accuracy (CI [81.0, 83.5]; Figure 2a) using full spectral features.

**Figure 2:**
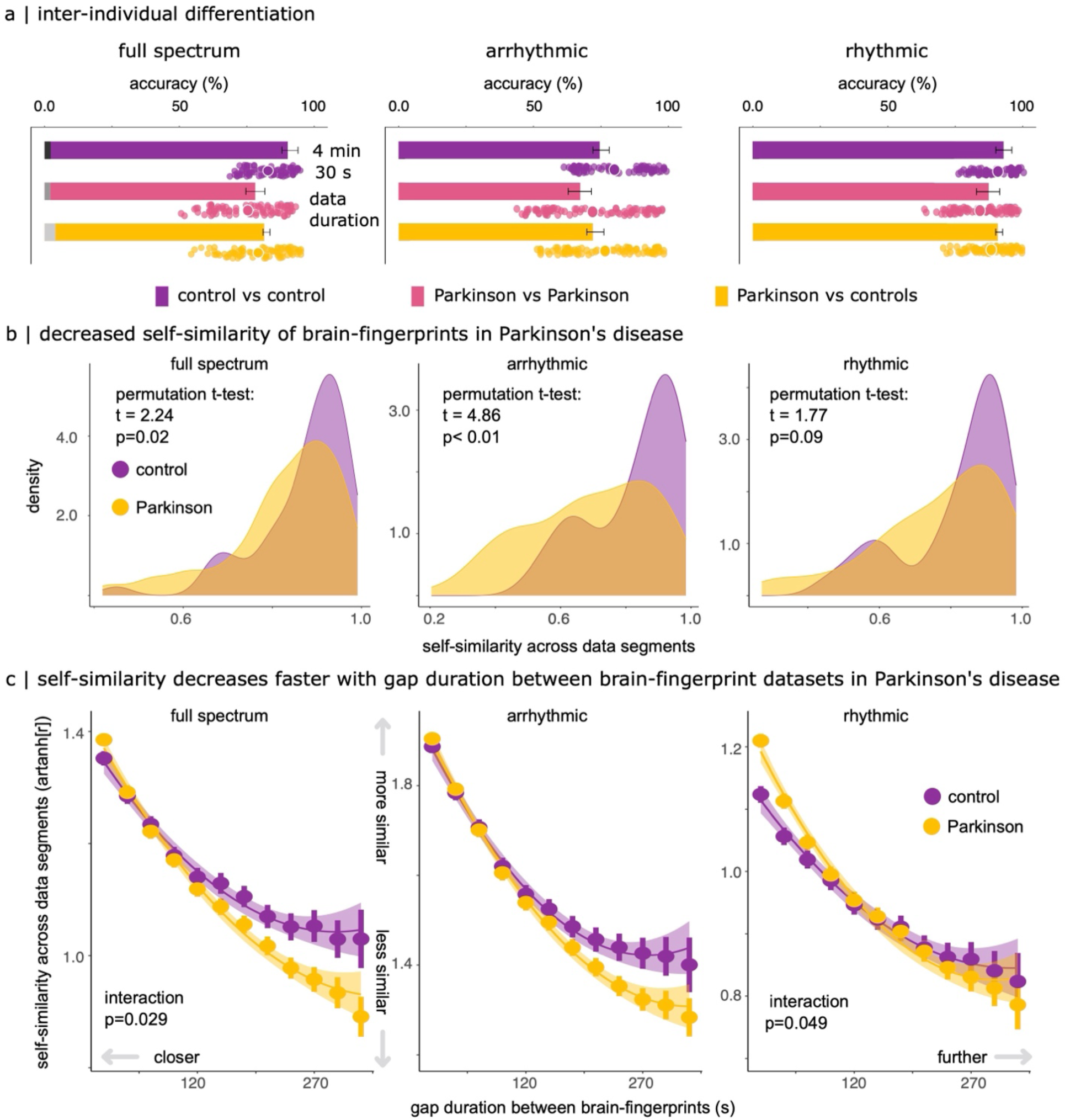
Differentiating Patients with Parkinson’s Disease from Healthy Controls Using Spectral Brain-Fingerprints. (a) Accuracy in distinguishing participants from their brain-fingerprints derived from full, arrhythmic, and rhythmic neurophysiological power spectra, estimated from 4-minute (bar plots) and 30-second (scatter plots) data segments. Scatter plots indicate differentiation accuracy for all brain-fingerprint pairs derived from all possible contiguous 30-second segments derived from the original 4-minute recordings. Grey segments at the base of the bar plots indicate control differentiation performances based on empty-room MEG recordings collected during each participant’ visit (refer to Methods). Error bars represent bootstrapped 95% confidence intervals. (b) Self-similarity statistics within participants for full spectral, rhythmic, and arrhythmic brain-fingerprints. The plots show the empirical density of self-similarity statistics between two consecutive brain-fingerprints in control and PD cohorts, with the PD group showing a wider distribution, suggesting more variability in patients for full spectral and aperiodic features. (c) Self-similarity of brain-fingerprints from brief (30-second) brain data segments across full spectral, rhythmic, and arrhythmic features. PD patients show lower self-similarity with increased gap durations between recordings (y-intercept shift downwards). The self-similarity of patient full spectrum brain-fingerprints decreases more rapidly as the gap duration between recordings increases. In contrast, the self-similarity of patient brain-fingerprints from rhythmic components was more self-similar than controls at short gap durations, and became comparable at longer durations. Shaded regions indicate the standard error on the mean.

To assess the respective contributions to this inter-individual differentiation from *arrhythmic* versus *rhythmic* neurophysiology, we parametrized the regional power spectra of the cortical time series into aperiodic (broadband, scale-free 1/f) and periodic (band-limited, oscillatory) components and used these data to recompute brain-fingerprints. The accuracy of inter-individual differentiation based on *arrhythmic* brain-fingerprints decreased to 74.1% between healthy controls (CI [72.0, 78.0]), 66.5% between patients (CI [62.9, 71.4]), and 71.5% accuracy individual patients and healthy controls (CI [69.6, 75.9]; Figure S1 and Supplemental Information). In contrast, the accuracy of inter-individual differentiation based on *rhythmic* brain-fingerprints increased to 92.6% (CI [90.0, 96.0]) among healthy participants, 86.7% (CI [82.9, 91.4]) between patients, and 90.5% (CI [89.9, 92.4]) between individual patients and healthy controls (Figure 2a bar plots).

We then sought to determine whether the present participants could be similarly differentiated based on brief, 30-second segments, thereby replicating our previous observations in younger healthy participants with older healthy adults and patients.^24^. We observed a similar pattern of differentiation accuracies: differentiation between healthy participants reached 84.9% (computed 95% CI [83.1, 86.7]), 77.2% for between patients (95% CI [74.4, 79.9]), and 81.2% for between patients and healthy controls (95% CI [78.7, 83.7]) for full spectral features. These results demonstrate the robustness of the spectral brain-fingerprinting approach with respect to data length (scatter plots in Figure 2a). Both brain-fingerprints of the arrhythmic and rhythmic components derived from brief segments of 30 seconds exhibited similar patterns, with arrhythmic brain-fingerprints differentiating between patients with lower accuracy than healthy participants.

### Moment-to-moment arrhythmic fluctuations are increased in Parkinson’s disease

We aimed to understand why the accuracy of differentiating PD patients from healthy controls varied so significantly, both with full spectral brain-fingerprints (77.2% vs. 89.8% accuracy) and arrhythmic ones (66.5% vs. 74.1% accuracy). To do this, we compared the similarity of brain-fingerprints within each dataset (self-similarity) to the similarity with fingerprints from other participants (other-similarity). Our analysis revealed no significant difference in *other-similarity* among healthy participants and PD patients when comparing full spectral brain-fingerprints (see Figure S2). However, we noted a significant reduction in the *self-similarity* of the patients’ full spectral brain-fingerprints (t=2.24, p=0.02; permutation t-tests; Figure 2b).

To better understand this effect, we analyzed the impact of arrhythmic versus rhythmic neurophysiological spectral components on self-similarity. We observed that *arrhythmic* brain-fingerprints in patients demonstrated reduced self-similarity (t=4.86, p<0.01; permutation t-tests), unlike *rhythmic* brain-fingerprints (t=1.77, p=0.09; permutation t-tests; Figure 2b).

Further, we investigated if this discrepancy could be linked to the increased moment-to-moment variability in the brain activity of PD patients within the recording session (as detailed in Methods under ’Temporal variability of the PD brain-fingerprint’; see Table S2-S4 and Figure 2c). Using the shorter 30-second data segments, we discovered that for full spectrum brain-fingerprints, the self-similarity decreases more rapidly in patients than in healthy controls as the duration of the gap between the data segments increases (β=-3.77, SE=1.73, 95% CI [-7.16, -0.38], p= 0.029; detailed in Table S2). This pattern was not significant for arrhythmic brain-fingerprints (β=-3.67, SE=2.33, 95% CI [-8.25, 0.92], p= 0.117; Table S3) but showed a similar trend. Conversely, *rhythmic* brain-fingerprints revealed a different pattern: for shorter time gaps between data segments, the neurophysiological activity in patients with PD was more self-similar than that of controls, becoming comparable over longer durations (β=-3.33, SE=1.69, 95% CI [-6.65, -0.02], p=0.049; Table S4). Together, these results suggest that the decreased differentiation accuracy observed in PD is related to an increased moment-to-moment variability of *arrhythmic* brain-fingerprints in PD.

### The Parkinson’s brain-fingerprint indicates disease stages

Given the noted temporal stability of rhythmic neurophysiological features in patients with Parkinson’s Disease (PD), we calculated the intraclass correlation (ICC) scores for each cortical region to identify the most consistent neurophysiological features in the rhythmic spectral brain-fingerprints across individuals^21,39^. We found distinctive patterns of rhythmic neurophysiology in varying brain regions between healthy controls and PD patients. The highest ICC values were in frontal and medial cortical regions for healthy controls (Figure S3a), and in the right pre- and post-central regions for PD patients (Figure 3a and Figures S3b).

**Figure 3:**
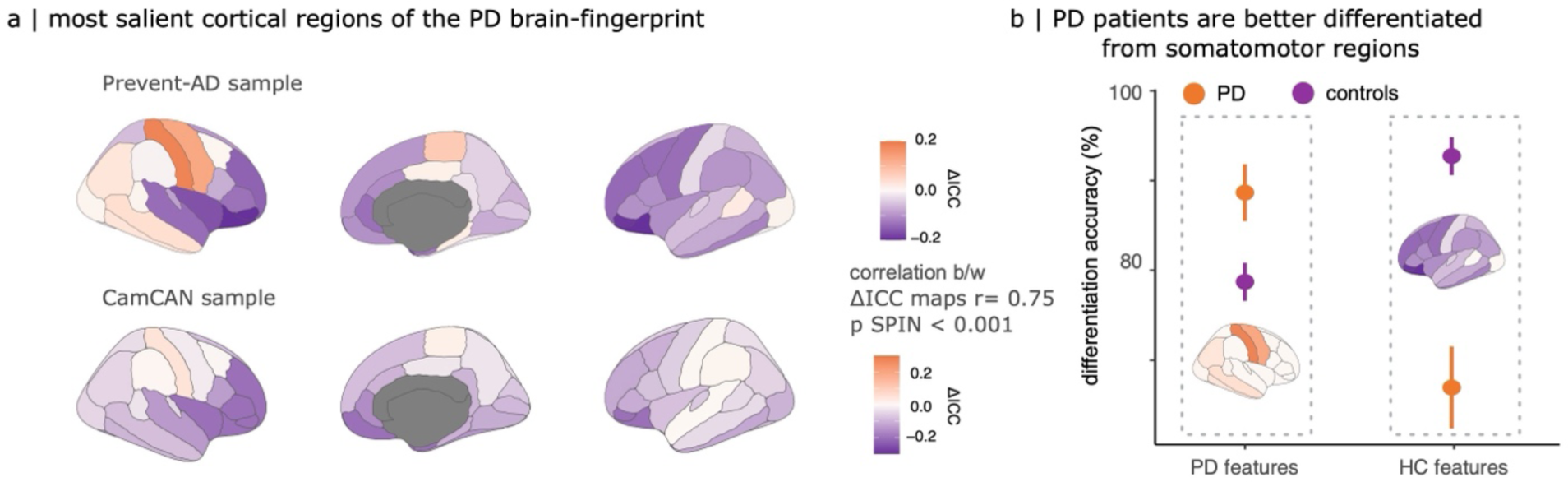
Comparative Analysis of Brain-Fingerprint Differentiation in Parkinson’s Disease and Control Groups. (a) Cortical maps comparing ICC scores for differentiating between patients and controls. Orange areas show regions where differentiation of individual patients is more effective than in controls. We replicated this finding in two independent samples of healthy controls: the Prevent-AD dataset (top panel) and the CamCAN dataset (bottom panel). (b) Differentiation accuracy from brain-fingerprints defined by top features for differentiating patients (left, cortical areas shown in orange) and top features for differentiating controls (right, cortical areas shown in blue).

We replicated these findings with an external sample of healthy age-matched controls from the Cambridge Center for Aging Neuroscience (CamCAN) dataset^40^ (Figure 3a, see Methods). We computed a cortical map of ICC values for the independent sample of healthy older controls and contrasted this map with the topography of patients with PD (i.e., ΔICC map). The cortical maps of the distinctive patterns of the PD brain-fingerprint obtained from using the two separate control samples were strongly correlated across control samples (r= 0.75, p< 0.001, p_spin_< 0.001).

To gauge how the spatial divergence between the rhythmic brain-fingerprints relates to individual differentiation, we created brain-fingerprints for both PD patients and healthy controls using the top 10% of ICC features specific to each group (Figure 3a). Utilizing the most distinctive features of the brain-fingerprints of PD patients, we achieved a differentiation accuracy of 78.7% ([76.6, 80.8] CI; Figure 2a) among healthy participants, and 88.7% ([85.5, 91.8] CI; Figure 3b) among PD patients. In contrast, using the features most salient in healthy control brain-fingerprints, we differentiated healthy participants with 92.7% accuracy ([90.6, 94.9] CI; Figure 3b), and PD patients with only 66.9% accuracy ([62.4, 71.5] CI; Figure 3b).

We then explored if the rhythmic brain-fingerprint of a patient could be indicative of their clinical disease stage. For this purpose, we developed binary classifiers to decode the disease stage based on rhythmic spectral features at each cortical parcel (detailed in Methods under ’Decoding disease staging from brain-fingerprints’). We classified the disease stage as either “early” or “advanced” according to the patients’ scores on the Hoehn & Yahr clinical scale (HY< 2 and HY≥ 2, respectively)^41,42^.

The cortical map of regional decoding accuracies revealed that it is possible to distinguish early from advanced clinical stages, exceeding chance levels, through electrophysiological brain activity. The most notable brain regions enabling this decoding were the right post-central and left caudal middle frontal gyri, showing decoding accuracies of 69.6% and 68.8%, respectively (Figure 4a). This data-driven approach uncovered that in these specific regions, there is a suppression of faster brain activity above 15Hz and an increase in slower activity (6-9 Hz) in the more advanced disease stages (Figure 3b, right panel).

**Figure 4:**
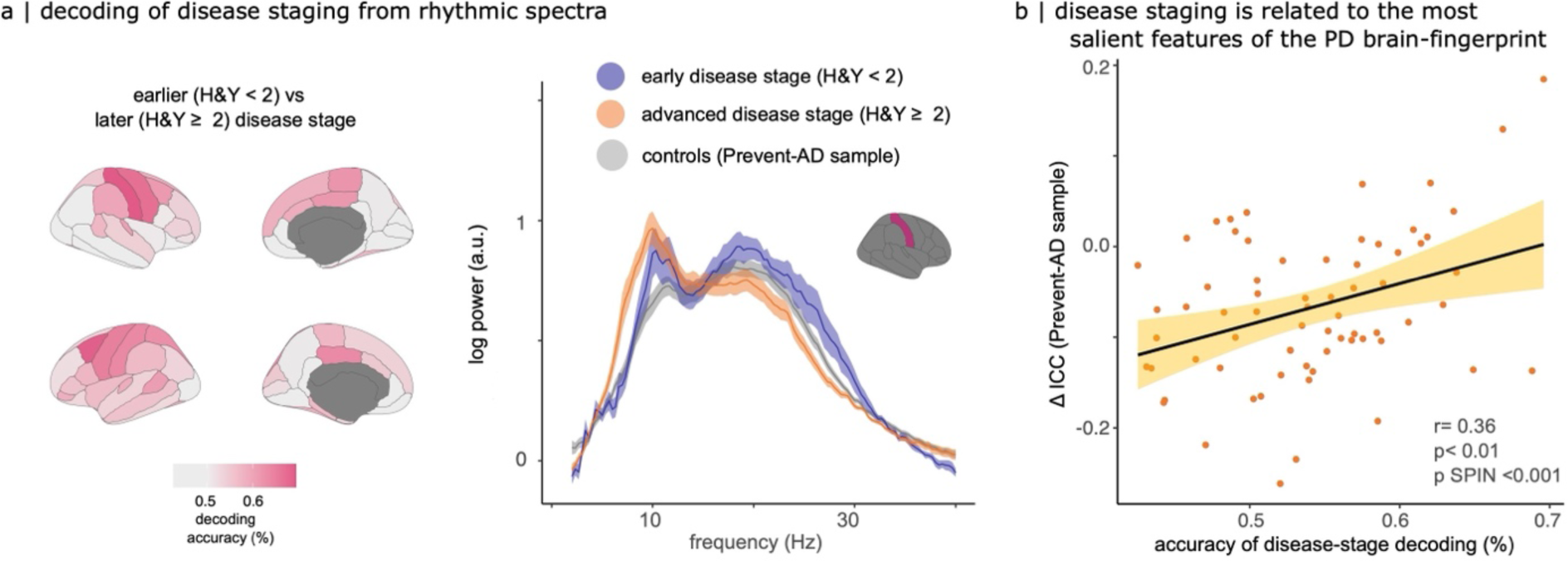
Decoding Stages of Parkinson’s Disease from Brain-Fingerprints. (a) Cortical topography of decoding accuracies for Parkinson’s disease stages (based on binarized Hoehn & Yahr scores). On the right, power spectra of resting-state neurophysiological activity in the right postcentral gyrus, the cortical region with the highest accuracy in disease stage decoding. Plots represent the average power spectrum for each group: healthy controls, early and advanced disease stages, with shaded areas indicating standard errors across groups. (b) Scatter plot showing how the decoding accuracy of Parkinson’s disease stages from brain-fingerprint features of each cortical parcel correlates with the saliency of each parcel, as determined by its ΔICC score.

Moreover, we found that the cortical map for disease-stage decoding aligns with the map of ICC difference scores (Figure 4b) and replicated this alignment using the CamCAN sample of healthy controls r= 0.36 (p< 0.01, p_spin_< 0.001) and r= 0.51 (p>0.001, p_spin_< 0.001), respectively. This consistency in findings was robust regardless of the cross-validation method employed for training the disease-stage classifiers (Figure S6).

### Aligning Parkinson’s Disease Brain-Fingerprints with Cortical Functional Gradients and Neurotransmitter Systems

We found that the regional disparities in prominent features of the rhythmic brain-fingerprint between Parkinson’s Disease (PD) patients and controls (indicated by ΔICC; see Figure 3a) were aligned with the unimodal-to-transmodal functional gradient of the cortical hierarchy^35^ (r=-0.49, p< 0.001, p_spin_<0.001; Figure 5a, with details in Methods). The most notable *rhythmic* brain-fingerprint features in healthy adults were associated with transmodal cortical regions. Conversely, the distinct features of the Parkinson *rhythmic* brain-fingerprint were more closely related to unimodal (i.e., primary sensorimotor) areas within the functional hierarchy of the cortex. Again, we replicated this effect using the CamCAN sample of healthy controls (r=-0.53, t*p*< 0.001, *p_spin_<0.001*; Figure S7).

**Figure 5:**
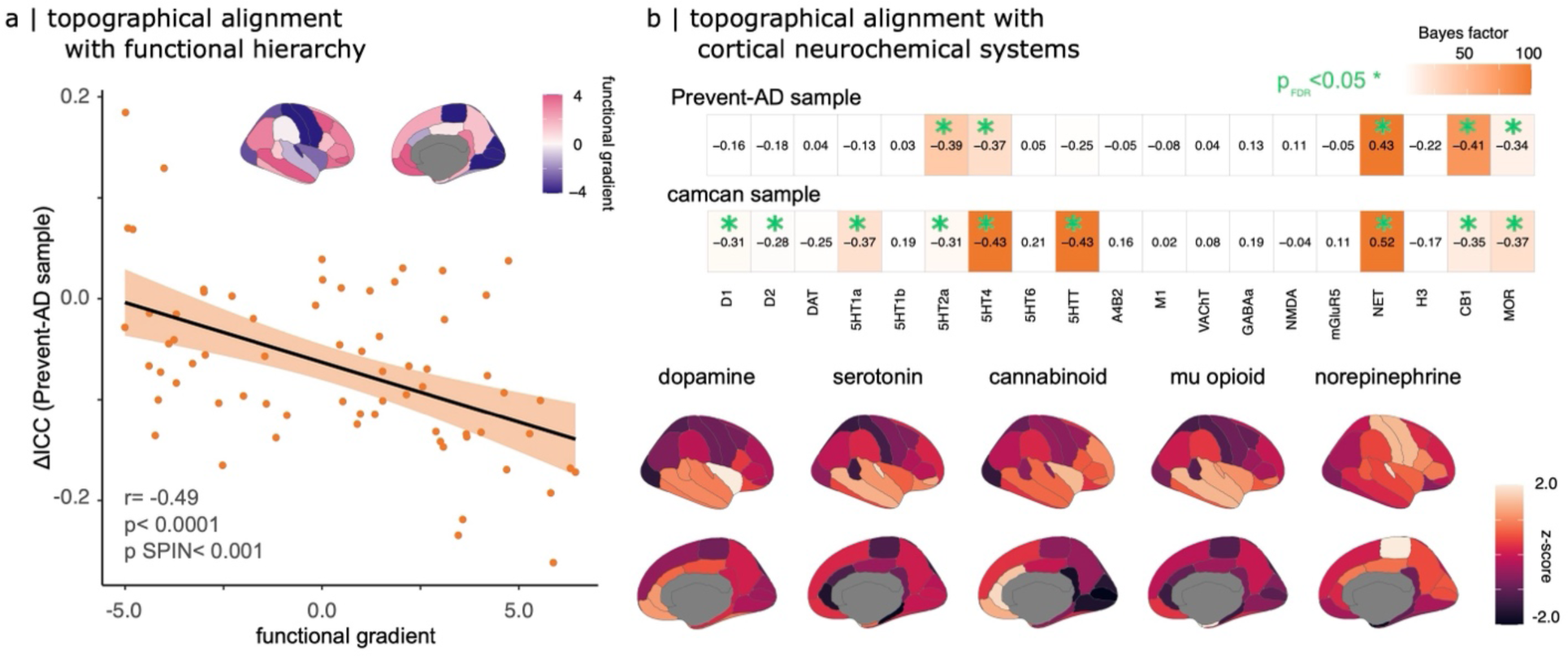
Correlation of Spectral Brain-Fingerprints with Cortical Functional Hierarchy and Neurotransmitter Systems. (a) Top: Cortical map illustrating the first unimodal-to-transmodal functional gradient, sourced from *neuromaps*^36^. Bottom: Linear association between the weights of cortical regions in this functional gradient (as per *neuromaps*) and their prominence in the PD brain-fingerprint (Figure 3a, top). (b) Top: Bayes factor analysis of the topographical alignment between PD brain-fingerprint features (from Figure 3a) and atlases of various cortical neurochemical systems, highlighting strong correlations particularly with serotonin, cannabinoid, mu-opioid, and norepinephrine systems. Each row represents data from different control samples. Bottom: Selected neurochemical cortical atlases, as obtained from *neuromaps*.

We further investigated if the most prominent features of the Parkinson’s brain-fingerprint were topographically related to the cortical distribution of major neurotransmitter systems. Using *neuromaps*^36^, we obtained 19 normative cortical maps representing 9 neurotransmitter systems (Figure 5b bottom) and assessed their spatial correlation with the cortical map of ICC difference scores (Figure 3a Prevent-AD sample; see Methods). Our analysis revealed significant correlations with several neurotransmitter systems, including serotonin-2a (r=-0.39, p_FDR_=0.006, p_spin_<0.001), serotonin-4 (r=-0.37, p_FDR_ =0.008, p_spin_ =0.008), cannabinoid-1 (r= -0.41, p_FDR_ =0.00045, p_spin_<0.001), mu-opioid (r=-0.34, p_FDR_ =0.018, p_spin_ =0.007) receptors, and the norepinephrine transporter (r=0.43, p_FDR_ =0.0040, p_spin_ <0.001). Notably, the cannabinoid, opioid, and serotonin systems, concentrated in temporal and frontal cortical regions, and corresponded with the most salient *rhythmic* brain-fingerprint features in healthy controls (Figure 5b & Figure S3a). Conversely, the pronounced presence of norepinephrine transporters in the somato-motor cortices mirrored the significance of *rhythmic* neurophysiology in these areas in PD patients (Figure 5b & Figure 3a).

This effect replicated using the CamCAN sample of healthy controls (Figure 3a). We found alignments with the cortical distributions of serotonin-2a (r=-0.31, p_FDR_ =0.02, p_spin_ =0.005), serotonin-4 (r=-0.43, p_FDR_ =0.002, p_spin_ =0.002), cannabinoid-1 (r= -0.35, p_FDR_ =0.01, p_spin_ =0.003), mu-opioid (r=-0.37, p_FDR_ =0.007, p_spin_ =0.002) receptors, and the norepinephrine transporter (r=0.52, p_FDR_ <0.001, p_spin_<0.001). Additionally, we observed a correspondence with the dopamine-1 (r=-0.31, p_FDR_ =0.02, p_spin_ =0.005), dopamine-2 (r=-0.28, p_FDR_ =0.04, p_spin_ =0.035), and serotonin transporter maps (r=-0.43, p_FDR_ =0.001, p_spin_ =0.003; Figure 5b).

### Robustness of Spectral Brain-Fingerprints Against Environmental and Physiological Artifacts

To ensure the reliability of spectral brain-fingerprints, we tested their robustness against environmental and physiological artifacts. We first evaluated environmental factors, specifically those related to the recording conditions on different days. To that effect, we used empty-room MEG recordings conducted around each participant’s visit. By processing these recordings in a manner similar to the participant data and mapping them onto the participant’s cortical surfaces with the same imaging procedure used for their MEG data, we established that environmental factors did not significantly contribute to individual differentiation. Notably, the differentiation accuracy based on these empty-room recordings was substantially lower than that achieved with actual spectral brain-fingerprints (<5%; see Figure 2a & Figure S1).

Further, we evaluated the influence of common physiological artifacts in MEG recordings, such as head motion, heart-rate variability, and eye blinks, on brain-fingerprinting. Our findings indicated that inter-individual differentiability was not significantly affected by cardiac or ocular artifacts (r= -0.04, p= 0.71 and r= -0.08, p= 0.46, respectively). However, there was a modest association with head movements in the PD cohort (r= 0.24, p= 0.04; Bayesian post-hoc analysis BF= 2.04; Figure S5). Consequently, we included head motion as a nuisance covariate in all subsequent regression analyses (detailed in Methods). We note that there were no significant differences in physiological artifact profiles between healthy controls and PD patients (head motion: t(64.34)= 0.41, p= 0.68; EOG: t(123.88)= -0.91, p=0.36; ECG: t(64.41)= -1.24, p=0.22).

Lastly, considering previous reports of cortical thickness abnormalities in PD^4–7^, we investigated whether these structural changes could partly explain the differentiability of PD patients from healthy controls. We derived cortical thickness measures from the structural MRI data of both groups, when available (n=134; Figure S4a). We standardized the patients’ cortical thickness maps using z-score transforms based on healthy controls. Our analysis revealed no significant linear relationship between individual differentiability and the average standardized cortical thickness in PD patients (b= -0.03, SE= 0.07, 95% CI [ -0.16, 0.11], p= 0.69; Figure S4b). Additionally, the cortical topography of the most salient Parkinson’s brain-fingerprint features did not align with the cortical thickness changes observed in patients (Pearson’s correlation: r=0.04, t(66)=0.34, p=0.73, p_spin_ =0.36). Thus, we conclude that the individual differentiability observed in PD patients based on their spectral brain-fingerprints is not significantly influenced by cortical thickness alterations associated with the disease.

## Discussion

Our study demonstrates the application and relevance of brain fingerprinting to Parkinson’s disease (PD) research. We derived brain-fingerprints from task-free MEG recordings and first replicated the prior observation that the brain-fingerprints of patients with PD present increased variability over short periods of time compared to healthy controls^30,31^. We identified that this effect is due in large part to the enhanced temporal variability of the *arrhythmic* component of the neurophysiological brain activity of PD patients, making them less distinguishable from one another. However, we observed that PD patients can be accurately differentiated from each other and from healthy controls based on brain-fingerprints derived from the *rhythmic* components of their ongoing electrophysiological brain activity. We further show that the distinct features of these rhythmic fingerprints correlate with disease staging and align with neurochemical systems impacted in PD, underscoring the potential for targeting neuromodulation therapies based on rhythmic cortical neurophysiology in PD.

### Alterations of Cortical Signaling in Parkinson’s Disease

Previous studies highlighted frequency-specific signaling abnormalities in PD, particularly in motor and subcortical structures^43,44^. Our findings align with this literature^10,45,46^, showing that the most distinctive brain-fingerprint features in PD patients localize to the primary sensorimotor cortex (Figure 3b left panel & Figure S3c), which correlates with their disease stages (Figure 3b). In particular, we found evidence of a link between atypical beta and theta band activities in the postcentral gyrus and disease stages. This aligns with previous findings linking beta-bursting in the motor network and sensorimotor cortex with symptom severity and treatment response to medication^8,10^ and deep brain stimulation of the subthalamic nucleus^47^.

The role of midline theta-band activity in PD^48–51^, thought to reflect cognitive processes^52,53^ and dopaminergic signaling^54,55^, was also confirmed in our study (Figure 4a). These findings are supported by previous research on theta neurostimulation’s effectiveness in alleviating motor symptoms, including when targeting the precentral gyrus^56–58^.

### Functional Decoupling of the Default Mode Network in Parkinson’s Disease

We observed that the most salient brain-fingerprint features of healthy controls align with regions of the default-mode network (DMN; Figure 5a & Figure S3a). Prior studies have noted functional decoupling of the DMN in PD during rest and task-based activities^59–62^, often linked to the dopaminergic system^59–62^. Yet, our data from patients on stable antiparkinsonian medication regimens may have moderated the saliency of DMN regions in the patients’ brain-fingerprints (Figure 3). Thus, our observation that the DMN, transmodal brain regions of the functional hierarchy do not contribute substantially to the Parkinson brain-fingerprint (Figure 5a) may reflect a normalization effect of medications^63,64^. These observations prompt further investigation into how responsiveness to medications relates to brain-fingerprints in transmodal brain regions.

### Neurochemical correlates of the PD brain-fingerprint

Our data suggest that monoamine neurotransmitters are closely associated with the brain-fingerprint of PD (Figure 5b). Specifically, we found that the cortical topography of serotonin 2a and 4 receptor densities is inversely related to the PD brain-fingerprint, while there is a direct association with the norepinephrine transporter.

This finding is in agreement with previous observations of the degradation of monoamines in PD^65^. We report negative relationships between the brain-fingerprint of PD and dopamine systems (Figure 3a). This effect was weak and inconsistent, possibly because changes in dopaminergic signalling caused by PD may primarily affect subcortical structures^66^ rather than the cortex, where our analyses were restricted. Further, the normative neurochemical system maps available in *neuromaps* were derived from an independent sample of adults, who were younger in age than the PD patients and aged-matched controls of the present study. Future research should explore these effects across subcortical structures and with normative atlases of neurochemical systems in older adults.

We also observed a negative alignment of the Parkinson brain-fingerprint with the cannabinoid receptor-1 (CB1) system (Figure 5b), supporting prior research that documents elevated CB1 receptor concentrations in PD^61^, and highlighting CB1 as a potential therapeutic target in PD^69^. Our present results also highlight the potential participation of the cannabinoid system in the neuropathophysiology of PD and encourage more research in this area.

### Enhanced Temporal Fluctuation of Arrhythmic Brain Activity in Parkinson’s Disease

Our study revealed that the brain-fingerprints of patients with Parkinson’s disease fluctuate more over short time spans compared to age-matched healthy individuals. This finding aligns with the decreased accuracy observed originally in differentiating individuals within the patient group (Figures 2b &c).

We anticipated greater variability in PD brain activity based on previous fNIRS research, which suggested a correlation between symptom severity and hemodynamic signal variability^31^. Additionally, studies using fMRI connectome brain-fingerprinting indicated reduced self-similarity in individuals at risk of or with mental health disorders^28,29^ and in PD patients^30^. Our data extend these findings to electrophysiology, pointing at increased within-subject variability of *arrhythmic* brain activity in PD as a possible source of such variability. We noted that differentiation accuracy using full spectral and arrhythmic brain-fingerprints in PD patients was lower compared to *rhythmic* brain-fingerprints, which achieved similar differentiation as seen in healthy controls (Figure 2a).

Recent research has linked alterations in PD patients’ arrhythmic brain activity to symptom severity^15,70,71^. Preliminary studies further suggest that baseline arrhythmic activity in the subthalamic nucleus may predict responses to neuro-stimulation protocols^13,14^. While these studies focused on group-level mean differences, our findings emphasize the significance of within-patient variability of arrhythmic brain activity in understanding individual disease manifestations. We hope our results promote further research into optimizing personalized rhythmic stimulation protocols for PD management by normalizing cortical dynamics.

Previous studies have also documented increased intra-individual variability in cognitive task performance in PD^72–74^, correlating with cognitive symptom severity^49,72,73^. The biological basis of this behavioral variability increase remains poorly understood^75^. fMRI research has linked moment-to-moment brain activity variability with cognitive performance^76–78^, and recent studies have related BOLD signal variability to the arrhythmic components of electrophysiology^79^. Consequently, we hypothesize that the heightened variability in PD behavioral markers may be associated with the observed increased temporal variability in arrhythmic brain activity.

The arrhythmic and rhythmic components of the neurophysiological spectrum indicate distinct neural mechanisms^33,34,80^. The arrhythmic spectrum’s slope is conceived as reflecting the balance of neuronal excitation versus inhibition^33,34^. Therefore, our findings tentatively suggest more fluctuant dynamics in cortical excitability in PD. This construct is in line with emerging insights that dynamics of spectral aperiodic components are key to understanding healthy aging and behaviors^80^.

### Potential Clinical Impact of Brain-Fingerprinting in Personalized Neuromodulation Therapies

The clinical utility of brain-fingerprinting hinges on its capacity to refine patient stratification, reveal novel disease characteristics, and inspire new treatment strategies.

Our findings demonstrate that brief brain recordings can distinguish individuals^24^, including those with Parkinson’s disease. We highlight the consistent within-participant stability of rhythmic brain-fingerprints in both patients and healthy controls (see Figure 2c), offering a unique insight into individual-specific brain activity. This consistency aligns with prior research showing the stabilization of spectral content in resting-state brain activity within 30 to 120 seconds of MEG recording^81^. This rapid stabilization is especially beneficial for clinical applications, particularly for patients with cognitive or motor impairments who may find longer recording sessions challenging.

The present study also suggests that personalized neuromodulation therapies should primarily concentrate on *rhythmic* neurophysiology, the most consistent electrophysiological characteristic within individuals, reflective of each patient and related to disease traits. Specifically, theta- and beta-frequency rhythms in the fronto-motor cortices emerge as potential prime targets for neurostimulation protocols aimed at normalizing disease-related neurophysiological changes^14,47,56,82^. Conversely, *arrhythmic* neurophysiological activity displayed less stability and individual specificity in Parkinson’s disease patients (Figure 2). This suggests that tracking the longitudinal variability of arrhythmic brain-fingerprints could enhance the definition and understanding of patient trajectories, as indicated here in Figure 2c. Incorporating this variability into adaptive neurostimulation therapies could potentially enhance clinical outcomes in Parkinson’s disease by reducing moment-to-moment fluctuations of core dynamics of brain activity related to the disease.

We also highlight the need to account for increased intra-individual variability of brain activity in disease states when developing statistical and machine learning models for disease classification. This consideration is crucial to ensure the scalability and generalizability of patient stratification methods.

In conclusion, our study underscores the clinical significance of brain-fingerprinting based on rapid neurophysiological activity dynamics. It sheds light on the clinical aspects of Parkinson’s disease, identifying specific brain regions and rhythms where disease impacts neurophysiological stability. We anticipate these insights will catalyze further research in population neuroscience and the development of personalized neuromodulation therapies for Parkinson’s disease and other neurodegenerative conditions.

## Data availability

The data are available through the Clinical Biospecimen Imaging and Genetic (C-BIG) repository (https://www.mcgill.ca/neuro/open-science/c-big-repository)^84^, the PREVENT-AD open resource (https://openpreventad.loris.ca/ and https://registeredpreventad.loris.ca)^85^, and the OMEGA repository (https://www.mcgill.ca/bic/resources/omega)^86^. Normative neurotransmitter density data are available from *neuromaps* (https://github.com/netneurolab/neuromaps)^36^.

## Code availability

All in-house code used for data analysis and visualization is available on GitHub https://github.com/jasondsc/PDneuralfingerprinting.

## Acknowledgements

Data collection and sharing for this project was provided by the Quebec Parkinson Network (QPN), the Pre-symptomatic Evaluation of Novel or Experimental Treatments for Alzheimer’s Disease (PREVENT-AD; release 6.0) program, and the Open MEG Archives (OMEGA). The funders had no role in study design, data collection and analysis, decision to publish, or preparation of the manuscript. The QPN is funded by a grant from Fonds de recherche du Québec-Santé (FRQS). PREVENT-AD was launched in 2011 as a $13.5 million, 7-year public-private partnership using funds provided by McGill University, the FRQS, an unrestricted research grant from Pfizer Canada, the Levesque Foundation, the Douglas Hospital Research Centre and Foundation, the Government of Canada, and the Canada Fund for Innovation. Private sector contributions are facilitated by the Development Office of the McGill University Faculty of Medicine and by the Douglas Hospital Research Centre Foundation (http://www.douglas.qc.ca/). The Brainstorm project is supported by funding to SB from the NIH (R01-EB026299-05). Further funding to SB for this study included Discovery grant from the Natural Science and Engineering Research Council of Canada (436355-13), and the CIHR Canada research Chair in Neural Dynamics of Brain Systems (CRC-2017-00311). This work was also supported by a grant F32-NS119375 (AIW) from the National Institutes of Health and a doctoral fellowship from NSERC (JDSC, JYH).

## Author contribution

JDSC contributed to the conceptualization of the project, curated data, formal analysis, methodology, software, visualization, and writing of the original draft. AW contributed to the conceptualization, curation of data, methodology, and reviewing & editing of the manuscript. JYH contributed to the software, methodology, and reviewing & editing of the manuscript. BM contributed to the methodology, supervision, and reviewing & editing of the manuscript. SB contributed to the conceptualization, methodology, supervision, funding acquisition, and reviewing & editing of the manuscript.

