## Supplemental Materials for "The neurophysiological brain-fingerprint of Parkinson’s disease"

### Supplemental Information

#### Participants.

The study participants were patients with Parkinson's disease from the QPN data repository and age-matched healthy controls from the Prevent-AD data repositories[1–3]. Table S1 provides their demographics, which we tested for sample differences in terms of age and education, using an unpaired t-test and gender and handedness with a Chi-square test. No significant differences were found. We replicated our analyses with a second sample of healthy controls obtained from the Cambridge enter for Aging Neuroscience (CamCAN) dataset (see Methods for demographics details).

Table S1: Participant demographics.

| | *Patients* | *Controls (Prevent-AD)* | *Uncorrected p-values* |
|---|---|---|---|
| *age* | 64.63 (8.66) | 61.98 (8.89) | 0.09 |
| *gender (female)* | 23 | 24 | 0.13 |
| *handedness (right)* | 67 | 48 | 1.0 |
| *education* | 15.11 (3.11) | 15.54 (3.58) | 0.48 |
| *Hoehn & Yahr score* | 1.97 (0.71) | NA | |
| *UPDRS III* | 32.55 (14.74) | NA | |
| *MoCA* | 24.43 (4.03) | NA | |

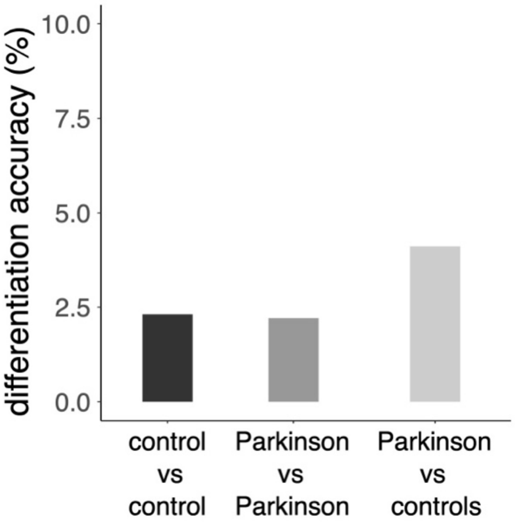


Figure S1: Differentiation accuracy from empty-room data.

Differentiation accuracies from brain-fingerprints derived from empty-room recordings performed around the MEG visit of each participant.

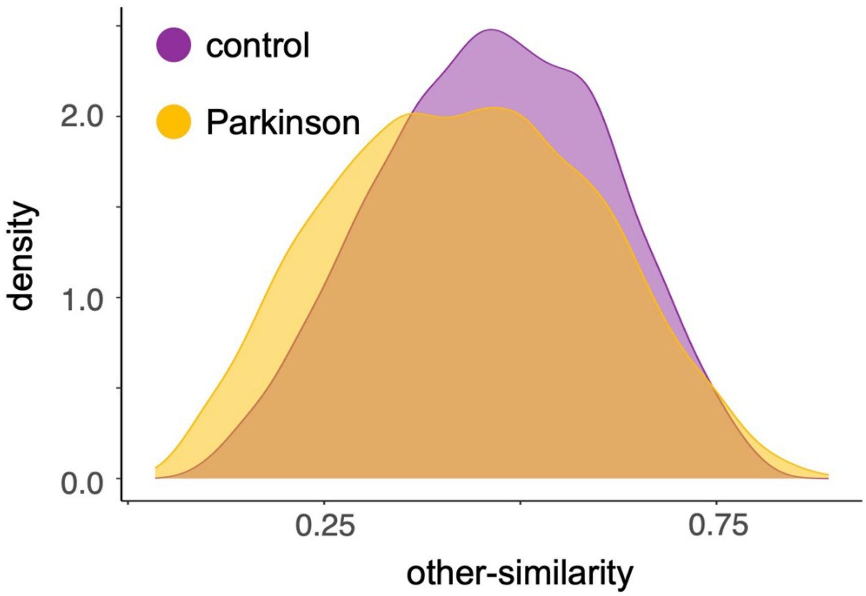


Figure S2: Empirical densities of *other-similarity* brain-fingerprint statistics

The empirical densities of inter-individual *other-similarity* brain-fingerprints statistics are similar between patients with Parkinson's disease and healthy age-matched controls. See **Moment-to-moment arrhythmic fluctuations are increased in Parkinson's disease** for a detailed description of the *self-similarity* statistics.

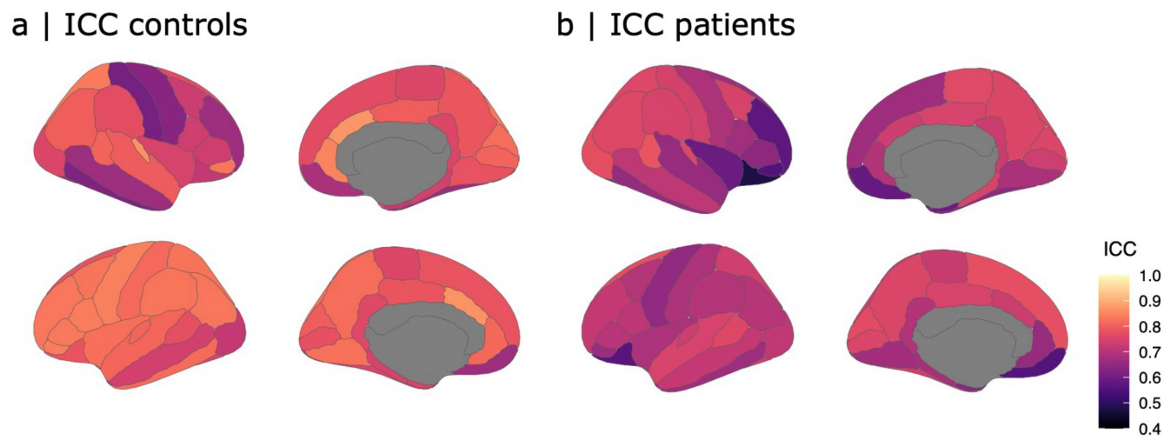


Figure S3: Relative contribution of rhythmic features for fingerprinting
Cortical maps of intra-class correlation coefficients (ICCs) for (a) controls (Prevent-AD), and (b) PD patients.

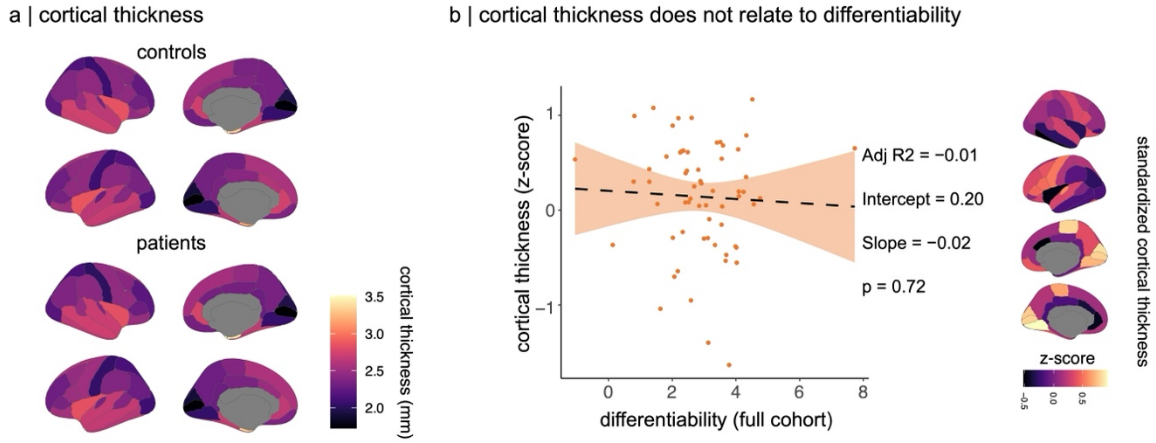


Figure S4: (a) Average cortical thickness measured in control participants and patients with PD. (b) left panel: linear regression analysis showing no relationship between individual differentiability and the average standardized cortical thickness of each patient (averaged across ROIs) (see Methods). Right panel: brain maps of average standardized cortical thickness of patients with PD.

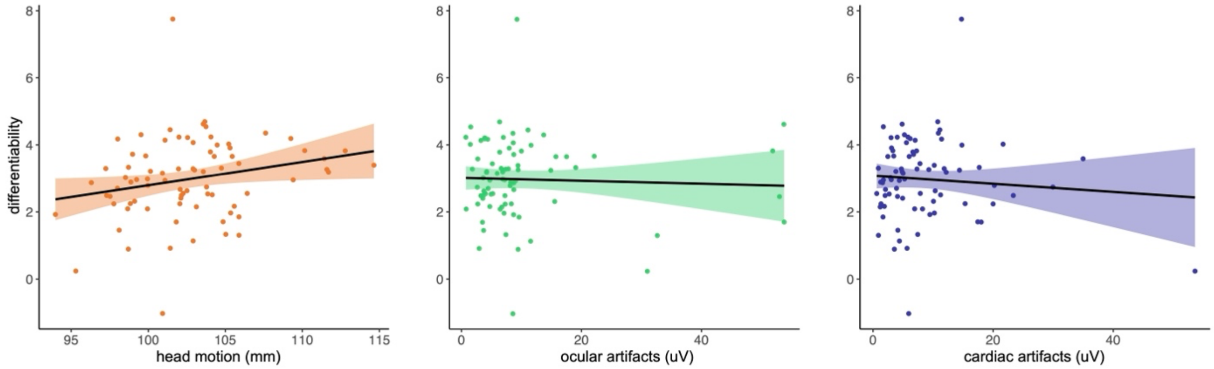


Figure S5: Individual differentiability does not relate to cardiac or ocular artifacts (middle and right panels). A moderate linear relationship exists between individual differentiability and head motion in the patient group.

Differentiability between patients with PD was correlated to head motion (r= 0.23, *p*= 0.04), but not to cardiac or ocular artifacts (r= -0.04, *p*= 0.71; r= -0.08, *p*= 0.46 respectively). There was, however, little evidence in favour of the relationship between head motion and differentiability (BF= 2.04; Figure S5).

#### Robustness and Replicability of Disease-Stage Decoding Using Spectral Brain-Fingerprints

The decoding of disease stages is related to the key features of the brain's spectral fingerprint. Specifically, the ability to distinguish disease stages based on regional brain-fingerprint characteristics correlates with changes in interclass correlation (ICC) between patients and control groups, as shown in Figure 3a.

To ensure that our findings were robust and not biased by our initial 80-20 cross-validation strategy, we conducted additional tests using 90-10 and 70-30 cross-validation ratios (i.e., training the decoder with 90% and 70% of the data respectively, and testing it with the remaining 10% and 30%). These tests yielded results consistent with our initial findings, as detailed in Figure S6.

Further, we replicated this analysis using the control sample from the Cambridge Center for Aging Neuroscience (CamCAN). The results confirmed that, similar to the Prevent-AD sample of healthy controls, the pattern of disease-stage decoding aligns with the cortical map of ICC difference scores. This was true for both 90-10 (correlation coefficient $r = 0.46$, $p < 0.01$, $p_{spin} < 0.001$) and 70-30 cross-validation ratios ($r = 0.54$, $p < 0.01$, $p_{spin} < 0.001$).

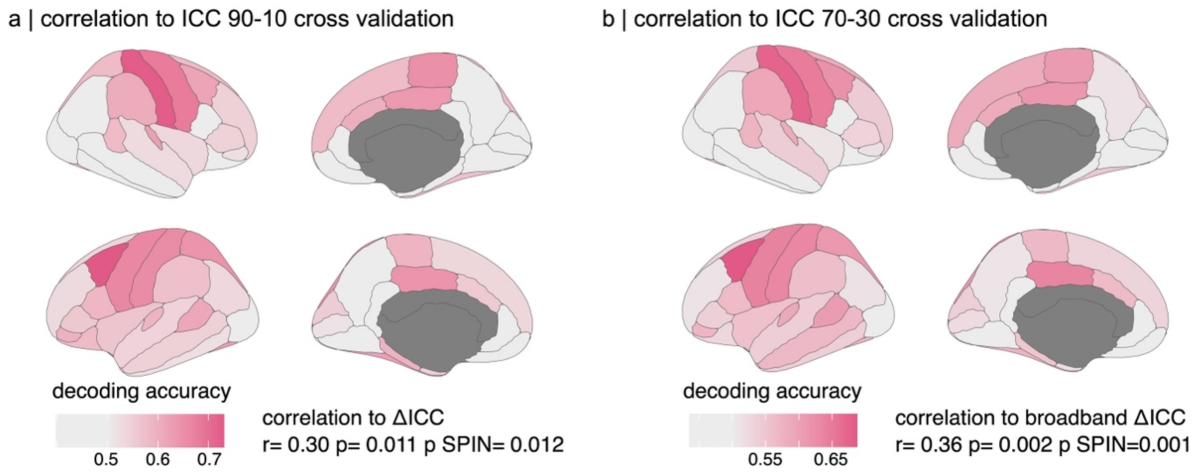


##### Figure S6: Stability of Disease Stage Decoding Across Various Cross-Validation Schemes

Decoding accuracies for Parkinson's disease stages (Hoehn & Yahr stages[5-6], binarized) derived from the rhythmic brain-fingerprint features from each cortical parcels of the Desikan-Killiany atlas using a) 90-10 and b) 70-30 cross-validation ratios. The cortical maps show consistency in the topographical distribution of decoding accuracies, irrespective of the cross-validation protocol employed. Moreover, a correlation persists between the decoding accuracy for Parkinson's disease stages at each cortical parcel and the prominence of the respective brain-fingerprint features within those parcels (Δ ICC; see Figure 3a), across different cross-validation strategies. These phenomena are further validated in the CamCAN cohort, as detailed in the preceding text.

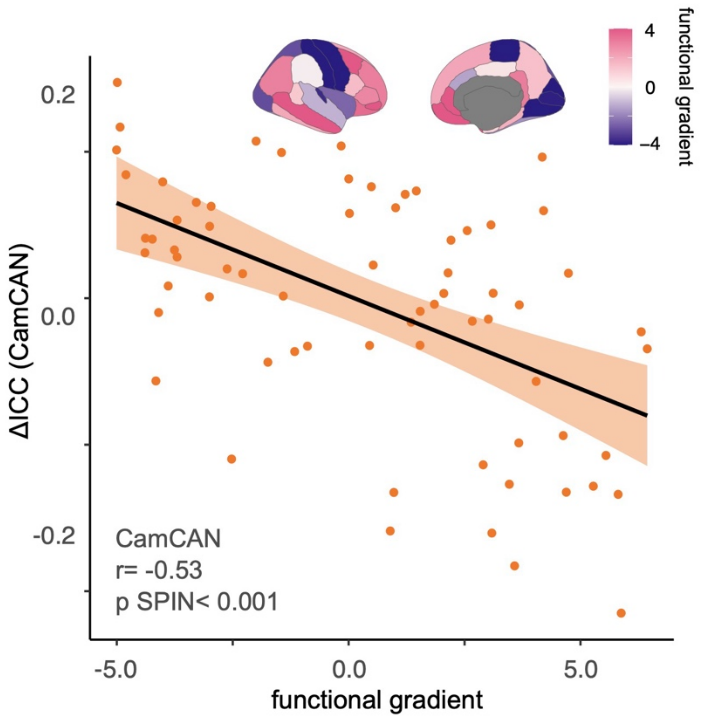


Figure S7: Topographical alignment of spectral brain-fingerprints with the functional hierarchy: CamCAN sample. Top: Cortical map illustrating the first unimodal-to-transmodal functional gradient, sourced from *neuromaps*[4]. Bottom: Linear association between the weights of cortical regions in this functional gradient (as per *neuromaps*) and their prominence in the PD brain-fingerprint (Figure 3a, bottom; CamCAN sample).

Table S2: Short-Term Variability in Self-Similarity of Brain-Fingerprints from 30-Second Datasets in Patients

| | atanh(self-similarity) | | |
|---|---|---|---|
| *Predictors* | *Estimates* | *CI* | *p* |
| (Intercept) | 1.19 | 1.13 – 1.25 | **<0.001** |
| Head motion | -0.03 | -0.06 – 0.01 | 0.149 |
| Group [Parkinson] | -0.02 | -0.10 – 0.05 | 0.546 |
| gap duration [ploy 1st degree] | -9.95 | -12.56 – -7.34 | **<0.001** |
| gap duration [ploy 2nd degree] | 2.76 | 1.68 – 3.83 | **<0.001** |
| Group [Parkinson] × gap duration [ploy 1st degree] | -3.77 | -7.16 – -0.38 | **0.029** |
| Group [Parkinson] × gap duration [ploy 2nd degree] | -0.03 | -1.43 – 1.36 | 0.961 |
| **Random Effects** | | | |
| $\sigma^2$ | 0.07 | | |
| $\tau_{00}$ SubjId | 0.05 | | |
| $\tau_{11}$ SubjId.poly(gap duration, 2)1 | 86.88 | | |
| $\tau_{11}$ SubjId.poly(gap duration, 2)2 | 7.32 | | |
| $\rho_{01}$ | 0.27 | | |
| | -0.40 | | |
| ICC | 0.46 | | |
| N SubjId | 133 | | |
| Observations | 10374 | | |
| Marginal $R^2$ / Conditional $R^2$ | 0.115 / 0.521 | | |

Table S3: Short-Term Variability in Self-Similarity of *arrhythmic* Brain-Fingerprints from 30-Second Datasets in Patients

| | atanh(arrhythmic self-similarity) | | |
|---|---|---|---|
| *Predictors* | *Estimates* | *CI* | *p* |
| (Intercept) | 1.63 | 1.57 – 1.70 | **<0.001** |
| Head motion | 0.02 | -0.02 – 0.05 | 0.359 |
| Group [Parkinson] | -0.02 | -0.11 – 0.06 | 0.608 |
| gap duration [ploy 1st degree] | -15.19 | -18.72 – -11.66 | **<0.001** |
| gap duration [ploy 2nd degree] | 4.82 | 3.29 – 6.36 | **<0.001** |
| Group [Parkinson] × gap duration [ploy 1st degree] | -3.67 | -8.25 – 0.92 | 0.117 |
| Group [Parkinson] × gap duration [ploy 2nd degree] | -0.47 | -2.47 – 1.52 | 0.641 |
| **Random Effects** | | | |
| $\sigma^2$ | 0.10 | | |
| $\tau_{00\ SubjId}$ | 0.06 | | |
| $\tau_{11\ SubjId.poly(gap\ duration,\ 2)1}$ | 161.32 | | |
| $\tau_{11\ SubjId.poly(gap\ duration,\ 2)2}$ | 19.28 | | |
| $\rho_{01}$ | 0.34 | | |
| | -0.46 | | |
| ICC | 0.42 | | |
| $N_{SubjId}$ | 133 | | |
| Observations | 10374 | | |
| Marginal $R^2$ / Conditional $R^2$ | 0.151 / 0.504 | | |

Table S4: Short-Term Variability in Self-Similarity of *rhythmic* Brain-Fingerprints from 30-Second Datasets in Patients

| | atanh(rhythmic self-similarity) | | |
|---|---|---|---|
| *Predictors* | *Estimates* | *CI* | *p* |
| (Intercept) | 0.98 | 0.92 – 1.05 | **<0.001** |
| Head motion | -0.03 | -0.07 – 0.01 | 0.147 |
| Group [Parkinson] | 0.02 | -0.07 – 0.11 | 0.646 |
| gap duration [ploy 1st degree] | -8.67 | -11.23 – -6.12 | **<0.001** |
| gap duration [ploy 2nd degree] | 2.08 | 1.02 – 3.14 | **<0.001** |
| Group [Parkinson] × gap duration [ploy 1st degree] | -3.33 | -6.65 – -0.02 | **0.049** |
| Group [Parkinson] × gap duration [ploy 2nd degree] | 1.17 | -0.20 – 2.55 | 0.095 |
| **Random Effects** | | | |
| $\sigma^2$ | 0.05 | | |
| $\tau_{00\ SubjId}$ | 0.06 | | |
| $\tau_{11\ SubjId.poly(gap\ duration,\ 2)1}$ | 85.02 | | |
| $\tau_{11\ SubjId.poly(gap\ duration,\ 2)2}$ | 9.14 | | |
| $\rho_{01}$ | 0.22 | | |
| | -0.29 | | |
| ICC | 0.58 | | |
| $N_{SubjId}$ | 133 | | |
| Observations | 10374 | | |
| Marginal $R^2$ / Conditional $R^2$ | 0.097 / 0.623 | | |
